# Pelvis perturbations in various directions while standing in staggered stance elicit concurrent responses in both the sagittal and frontal plane

**DOI:** 10.1101/2022.07.18.22277773

**Authors:** Michelle van Mierlo, Jean A. Ormiston, Mark Vlutters, Edwin H.F. van Asseldonk, Herman van der Kooij

## Abstract

Walking very slowly increases the time spent in the double support phase, which could be resembled by the staggered stance posture. Maintaining balance in this posture is important in order to continue walking safely. We therefore aimed to increase the understanding of balance recovery in staggered stance. We studied balance responses on joint- and muscle level to pelvis perturbations in various directions while standing in this posture. Ten healthy individuals participated in this study. We used one motor beside and one behind the participant to apply perturbations in mediolateral (ML), anteroposterior (AP) and diagonal directions, with a magnitude of 3, 6, 9 and 12% of the participant’s body weight. Meanwhile motion capture, ground reaction forces and moments, and electromyography of the muscles around the ankles and hips were recorded. The perturbations caused movements of the centre of mass (CoM) and centre of pressure (CoP) in the direction of the perturbation. Furthermore, these were often accompanied by motions in a direction different from the perturbation direction. After ML perturbations and diagonal perturbations transverse to the line between both feet, large and significant CoM and CoP deviations were present in the sagittal plane. Also, stronger responses on joint and muscle level were present after these perturbations, compared to AP and diagonal perturbations collinear with the line between both feet. The hip, knee and ankle joints significantly responded to all perturbation directions, but in different manners and modes of cooperation. To conclude, standing in a staggered stance posture makes individuals more vulnerable to perturbations in ML direction and transverse to the line between both feet, requiring larger responses on joint level as well as contributions in the sagittal plane.

## Introduction

Research into human biomechanics, and in particular human balance recovery after perturbations, is vital to the development of technologies supporting rehabilitation of individuals with movement disorders. One such technology is an exoskeleton emulating a natural human gait and stance behaviour. Most exoskeletons have a slow walking speed compared to normal gait. This increases the duration of the double support phase during the gait cycle while decreasing the frontal stability of the user of the exoskeleton [1, 2]. Successful shifting of the body weight from the trailing to the leading leg during the double support phase is crucial for maintaining balance. This may be jeopardized due to the prolonged duration of this phase when walking with an exoskeleton [3]. Balance control in the double support phase during slow walking can be approximated with a static staggered stance, in which an individual stands still with the leading and trailing foot a step length and step width apart [4]. Studying the responses to perturbations in this posture could improve the understanding of human balance control in a staggered stance posture.

The ability to maintain balance is largely dependent on the size and orientation of the base of support (BoS) [4, 5]. A larger BoS allows for a larger displacement of the centre of pressure (CoP). This is one of the mechanisms to control balance and shown to be effective in controlling the centre of mass (CoM) during double support [6, 7]. In case of a narrow BoS another strategy will mainly be used, being the counter-rotation mechanism, induced by movement of the upper body, changing the orientation of the GRF [6, 8]. Coordinated hip, knee and ankle joint moments can redirect the ground reaction force (GRF) and modulate the CoP position in order to control the (CoM) and whole body angular momentum [9]. The staggered stance posture, with a large anteroposterior (AP) BoS, will allow for an effective use of the ankle strategy in the sagittal plane and for a large AP weight shift. However, maintaining balance in the frontal plane might be more challenging, since there is limited space for CoP modulation and weight shift.

Staggered stance is a hybrid of tandem and parallel stance [10]. In parallel stance, where the feet are next to each other, unperturbed sagittal plane balance is dominated by modulations of the ankle moments. For the frontal plane hip ab- and adduction moments realize the major contribution to balance. Contrarily, during quiet tandem stance, in which one foot is in front of the other, the hips account for the dominant balance response in the sagittal plane through flexion and extension. Mediolateral (ML) stability during tandem stance is predominantly governed by ankle in- and eversion with a smaller contribution from the hip ab- and adduction [4]. Standing in a staggered stance posture combines various of the balance responses and sensitivities of the tandem and parallel stance [4, 10, 11]. It also allows for more possibilities to recover balance in both the sagittal and frontal plane, compared to the tandem or parallel stance, which often has stronger mechanical constraints in either of the planes [10]. A study of postural sway during unperturbed staggered stance shows that for balance in the sagittal plane the ankle joint must cancel out a destabilizing hip load/unload balance mechanism [4]. This is unlike the frontal plane, were the hip and ankle joint reinforce each other to maintain balance [4, 11, 12]. Overall, for a staggered stance posture with naturally the most weight on the trailing leg, individuals tend to use balance strategies more similar to those used in parallel stance [10], while the CoP variability increased in the ML direction [4, 11].

Extensive research has been done to analyze human balance recovery after balance perturbations such as: multi-directional surface translations [8, 13–15], external forces applied to the pelvis [12, 16–20], visual perturbations [11] and self induced perturbations [21], in parallel or tandem stance. These studies give insights into the use of the hip, knee and ankle joints in order to maintain balance. The studies by Henry et al. [13] and Matjacic et al. [16] showed that, while standing in a parallel stance, diagonal perturbations provoke the largest joint and muscle responses. This large response is caused by a combination of the responses to an AP and ML perturbation [13, 16]. Various studies also showed that responses in both the frontal and the sagittal plane can be observed after perturbations in a single plane [11, 13, 16, 21]. For example tibialis anterior and rectus femoris activity after ML perturbations and adductor longus activity after anterior perturbations [13]. Lee et al. [21] showed a dependency of the response on the stance posture as well [21]. After self induced backward perturbations they showed that individuals had larger ML CoP displacement while standing in staggered stance, compared to parallel stance [21]. These studies have shown a coupling between the frontal and sagittal plane. However, it is not clear yet how balance recovery strategies in multiple planes are used after perturbations from different directions, while standing in staggered stance.

In this study we aim to establish how hip, knee and ankle joints responses contribute and co-operate in balance recovery after pelvis perturbations in various directions while standing in a staggered stance. Since the size and orientation of the BoS largely affect the risk of losing balance, the staggered stance posture influences the sensitivity to certain perturbation directions. Individuals will be more vulnerable to perturbations in the direction where the BoS is the smallest, which is in the ML direction and in the direction transverse to the line between both feet. It is hypothesized that motions in the sagittal plane will play an important role after all perturbation directions. Because of the staggered stance posture there will be a coupling between the motions in both the sagittal and frontal plane, allowing for balance recovery contributions from the sagittal to the frontal plane and vice versa. Since the BoS in the sagittal plane is the largest, the joints can induce larger modulations in the CoP and horizontal GRF, facilitating an efficient recovery of the CoM.

## Materials and methods

### Participants

This research was approved by the local ethical committee and all participants signed an informed consent form in accordance with the Declaration of Helsinki before participating in the study. Ten participants, five female and five male, with no known history of neurological, muscular or orthopedic problems participated in this study. The participants had an average (±SD) age of 23.6 ± 2.9 years, height of 1.76 ± 0.05 m, leg length of 0.91 ± 0.06m measured from the ground to the trochanter major and weight of 69.8 ± 7.8 kg.

### Setup

The experiments were carried out on a split-belt treadmill (custom YMill, Motek medical, Culemborg, The Netherlands), with the belts standing still. Two force plates were present beneath the belts for the measurement of GRFs and moments. Two motors (SMH60, MOOG, Nieuw-Vennep, The Netherlands) were placed on the rear and side of the treadmill, see the top-down view in Fig 1a. The participants wore a modified universal hip abduction brace (Distrac Wellcare, Hoegaarden, Belgium; weight 1 kg) which was attached to the motors via horizontal carbon rods and a lever arm of 0.3 m. Load cells (Model LR350 FUTEK, Los Angeles, CA, USA) where positioned in the horizontal rods, to measure and control the applied forces. The motors were controlled via the main computer (Linux, Ubuntu 16.04 LTS) and seven secundary devices: four Beckhoff modules (three analog input and one analog output, Beckhoff Automation GmbH, Germany), Haptic control unit (Moog PC CB79047-401 HCU, Nieuw-Vennep, The Netherlands) and two motor drives (Moog MSD 3200 Servo Drive, Nieuw-Vennep, The Netherlands). An admittance controller was used to minimize the interaction forces during standing and to track the desired forces at the moment a perturbation was given. A detailed description of this controller can be found in [22]. A screen was positioned in front of the treadmill, which was used to give the participant feedback on the position of their CoM and feet as well as the desired CoM position (see section ‘Experimental protocol - Control position centre of mass’ for more details).

**Fig 1.**
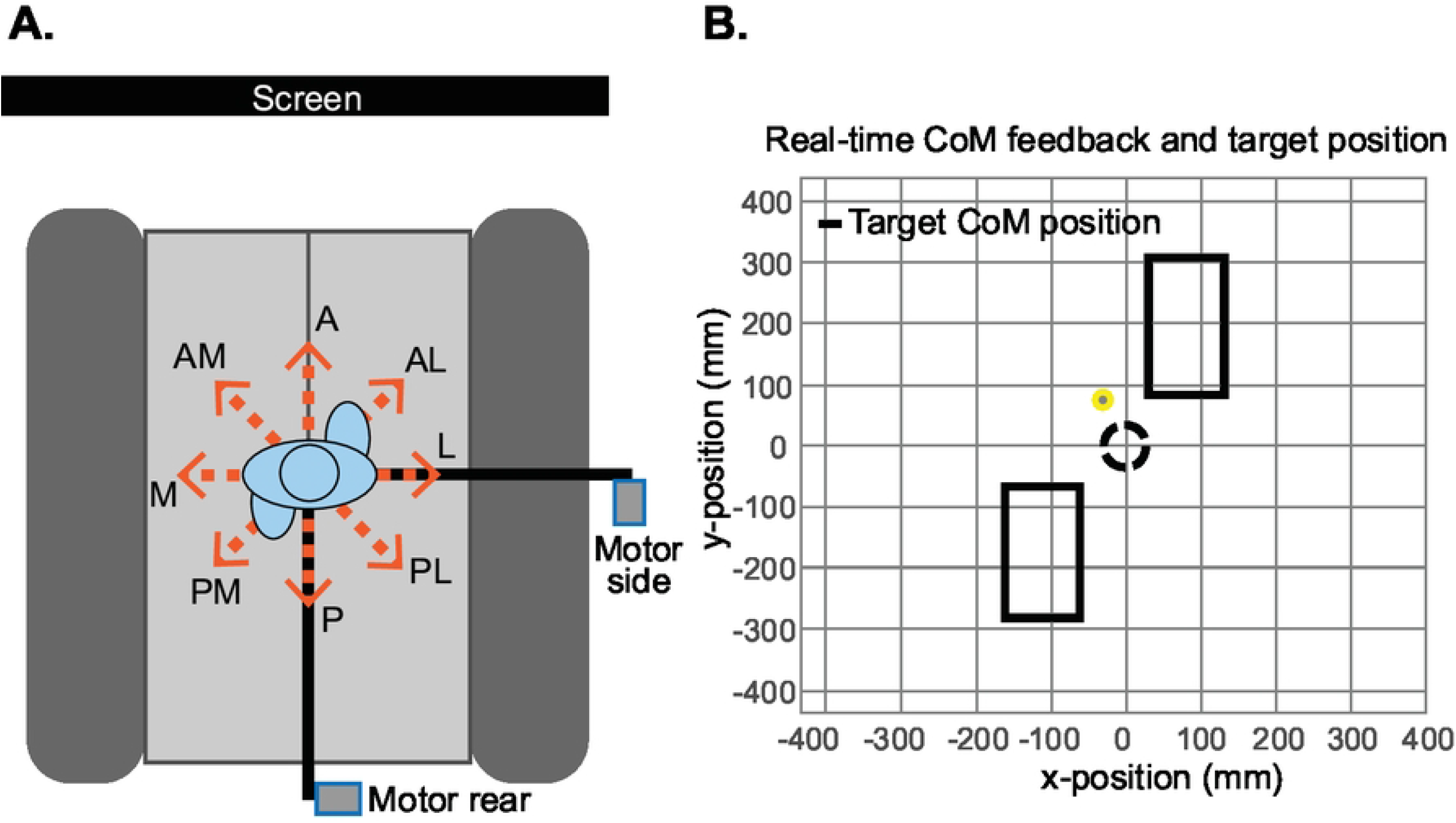
Experimental setup. A) Schematic top-down view of the setup, with one motor (side) placed to the right of the participant, and the other motor (rear) placed posterior to the participant. The orange arrows indicate the 8 perturbation directions. A = anterior, P = posterior, M = medial, L = lateral, AM = anterior-medial, AL = anterior-lateral, PM = posterior-medial, PL = posterior-lateral. A screen is placed directly in front of the participant displaying the position of their feet and CoM as well as the desired CoM position in real-time. B) Feedback presented on the screen. Top-down view of the participant’s feet, depicted with the rectangles and the yellow dot presenting the CoM. The black dashed circle is the target CoM position.

### Data collection

Kinematic marker data was acquired using an 8-camera infrared motion capture system (Oqus 600+, Qualysis, Götenborg, Sweden). The data was recorded at 128 Hz with the Qualysis Track Manager software (QTM, Qualysis, Götenborg, Sweden). In total six marker clusters were used on: the right and left shank and thigh, the sternum and on the front of the pelvis brace. Twenty-three individual markers were placed on bony landmarks using double-sided tape: on the 7^th^ cervical vertebra and the right and left calcaneus, 1^st^ and 5^th^ metatarsal heads, toes, medial and lateral malleoli, medial and lateral epicondyles of the femur, anterior and posterior superior iliac spine and acromia. The analog data measured by the force plates and EMG electrodes (Delsys Bagnoli, Natrick, USA) were recorded via an analog interface (Kistler 5695A DAQ) at 2048 Hz, synchronised with the motion capture data. Twelve wired surface EMG electrodes were placed according to the Seniam guidelines on the following muscles of the right and left leg: gluteus medius (mGMe), gluteus maximus (mGMa), adductor magnus (mAM), soleus (mSOL), tibialis anterior (mTA) and peroneus longus (mPL) [23]. The interaction forces between the participant and the motors were recorded at 1000 Hz, via the computer controlling the motors. This computer was also sending a synchronisation signal, which was recorded via the analog interface, to synchronise the kinematic data with the forces.

### Experimental protocol

#### Participant preparation

The reflective markers and marker clusters were attached to the participant, along with the wired EMG electrodes. Before starting the experiment the maximum voluntary contraction (MVC) was recorded, by performing a muscle-specific exercise for each individual muscle. The participant wore a safety harness (Honor, FBH-10) to prevent injury in case of a fall.

#### Staggered stance posture

During the experiment, participants stood still on the treadmill in a staggered stance, with the right foot in the leading position. The locations where the participant had to place their feet were marked on the treadmill. The step width and length were based on the average step width (0.15 m) and length (0.40 m) during walking at 0.5 m s^*−*1^ [24]. These measures were scaled with a factor determined by the participant’s leg length (*l*) with respect to the average leg length *l*_*av*_ = 0.91 *m* from Wu et al. [24]: *Scale factor* = *l/l*_*av*_. Participants were asked to stand up straight with the arms crossed over their torso to prevent contributions of arm swing to the balance recovery and to avoid collision between the participant’s arm and the rod on the side. Participants were also requested to stand with their knees slightly bent to prevent locking of the knee joint. They were instructed to refrain from stabilizing themselves by using the bars on the sides of the treadmill, unless this was really necessary and a side step did not suffice.

#### Control position centre of mass

To control the initial posture of the participant, perturbations were only given when the participant’s CoM was within a certain target position for at least 3 s. In order to assume this position, the participants received feedback of their CoM and feet position as well as the target CoM position via the screen in front of them. To generate this feedback the force plate and marker data were used in real time via a connection between the QTM SDK and a Python GUI. Since the participants were standing still, we used the CoP location as a representation of the particpants’s CoM. Fig 1b shows a screenshot of what the participants saw on the screen, representing a top-down view of the positions of the feet and CoM. The locations of the rectangles, representing the feet, were based on the position data of the 1^st^ and 5^th^ metatarsi together with the calcanei. The CoM target position approximates the halfway point during the double support phase, when the participants shift their weight from the trailing foot to the leading foot. A margin with a radius of 3 cm was taken around this point and displayed as a circle on the screen.

#### Perturbations

Pelvis perturbations were given in 8 different directions: anterior (A), posterior (P), medial (M), lateral (L) and diagonally anterior-medial (AM), anterior-lateral (AL), posterior-medial (PM) and posterior-lateral (PL), shown in Fig 1a. The perturbations were given at 4 different magnitudes (3%, 6%, 9% and 12% of the participant’s body weight) and lasted for 150 ms. The perturbations were given when the participant’s CoM was within the target for a random time between 3 and 5 s. Each participant performed 8 trials, containing each unique perturbation (direction and magnitude) once in a randomised order (8 × 4 = 32 perturbations per trial). In total this resulted in 256 perturbations per participant.

### Data processing

#### Pre-processing

Using the QTM software the recorded marker trajectories were labeled and all missing samples were filled with the polynomial gap filling tool. Further processing was done with Matlab (2022a, MathWorks). The marker and force plate data were filtered with a zero phase 4^th^ order 10 Hz low pass Butterworth filter. OpenSim 4.2 was used to scale the generic 23 segment model (gait2392) for each participant [25]. The inverse kinematics, analyze and inverse dynamics tools of OpenSim were used to obtain the joint torques, and positions and velocities of each segment and the total body. The EMG amplifier (Delsys) had a build in filter, filtering the data to a 20-450 Hz bandwith. We detrended the data by subtracting the mean. This was followed by a zero phase 1^st^ order 48-52 Hz Butterworth bandstop filter, rectification and zero phase 2^nd^ order 10 Hz Butterworth low-pass filter. The EMG data of each muscle was normalized to the maximum value of the corresponding muscle recorded during the MVC.

#### Data selection

As we were interested in assessing balance recovery strategies within the double support phase, all balance responses that involved taking a step were removed from the data. A step was identified when the following events were detected: 1) After a perturbation, for a duration of at least 0.04 s the vertical component of the GRF of one of the belts was lower than the threshold value of 20 N; 2) There was a change of at least 0.05 m of the toe and 5^th^ metatarsus marker of the corresponding foot.

#### Outcome measures

To indicate the balance sensitivity to the different perturbation magnitudes and directions, the number of steps for each perturbation condition were counted and expressed as the % of the total number of perturbations of the corresponding condition. To quantify the rate of the response, the time to the point of return was defined. This was expressed as the time from the instant the perturbation started until the CoM velocity in both the AP and ML directions was reversed into the direction of the starting position. A range of the first 1.5 s after the start of a perturbation was selected for the following outcome measures: CoP position, CoM position, EMG activity, and joint moments. To determine the maximum deviation of the CoP and CoM position for both the AP and ML directions, the largest deviation with respect to their starting position were considered within the 1.5 s window. For the EMG activity and joint moment outcome measures the mean value was taken over this range. Averages have been taken over the 8 repetitions of each perturbation condition (direction and magnitude) within each participant, followed by an average across all participants. Baseline measures were taken for the EMG activity and joint moments over 1 s before the start of the perturbation. For the baseline value averages have been taken over all repetitions within each participant, followed by an average across all participants.

### Statistics

The effect of the perturbations on the various outcome measures was assessed with linear mixed models. This analysis was performed in R4.2.0 (R Core Team, 2021, Vienna, Austria). For each perturbation direction separately, linear mixed models were fitted for the following outcome measures: maximum deviation of AP CoM, ML CoM, AP CoP and ML CoP, EMG of the mSOL, mTA, mPL, mGMa, mGMe and mAM of the leading (right) and trailing (left) leg, and the joint moments of the ankle in-/eversion and plantar-/dorsiflexion, knee flexion/extension, hip flexion/extension and ab-/adduction and lumbar bending and extension. The perturbation magnitudes were added as a fixed effect and for the intercept and slope, random effects were included to take into account the participant effects. The main effects were tested with a significance level of α = 0.05 using the Wald t-test with a Kenward-Roger correction for the degrees of freedom.

## Results

### Effect perturbations

The staggered stance posture resulted in a weight distribution of 60.3% on the trailing leg. After strong perturbations transverse to the line between both feet (this will further be used to refer to the following perturbations: M, L, AM and PL), participants had to take a step to recover balance more often compared to the perturbations approaching collinearity with the line between both feet (further used for the A, P, AL and PM perturbations). For the 12% magnitude perturbations this was sometimes in more than 60% of the perturbations, Fig 2a. Therefore this perturbation magnitude was removed from the rest of the analysis. In addition, the analysis of the time to the point of return demonstrated that the participants were challenged most by the perturbations transverse to the line between both feet. The time to the point of return increased with the perturbation magnitude, up to 0.8 s for the strongest perturbations in these directions, Fig 2b. This effect was less present for the perturbations collinear with the line between both feet, for which the time to the point of return stayed around 0.4-0.5 s. For all perturbations it holds that after the the end of the perturbation it took some extra time before the CoM started to move in the direction of the starting position

**Fig 2.**
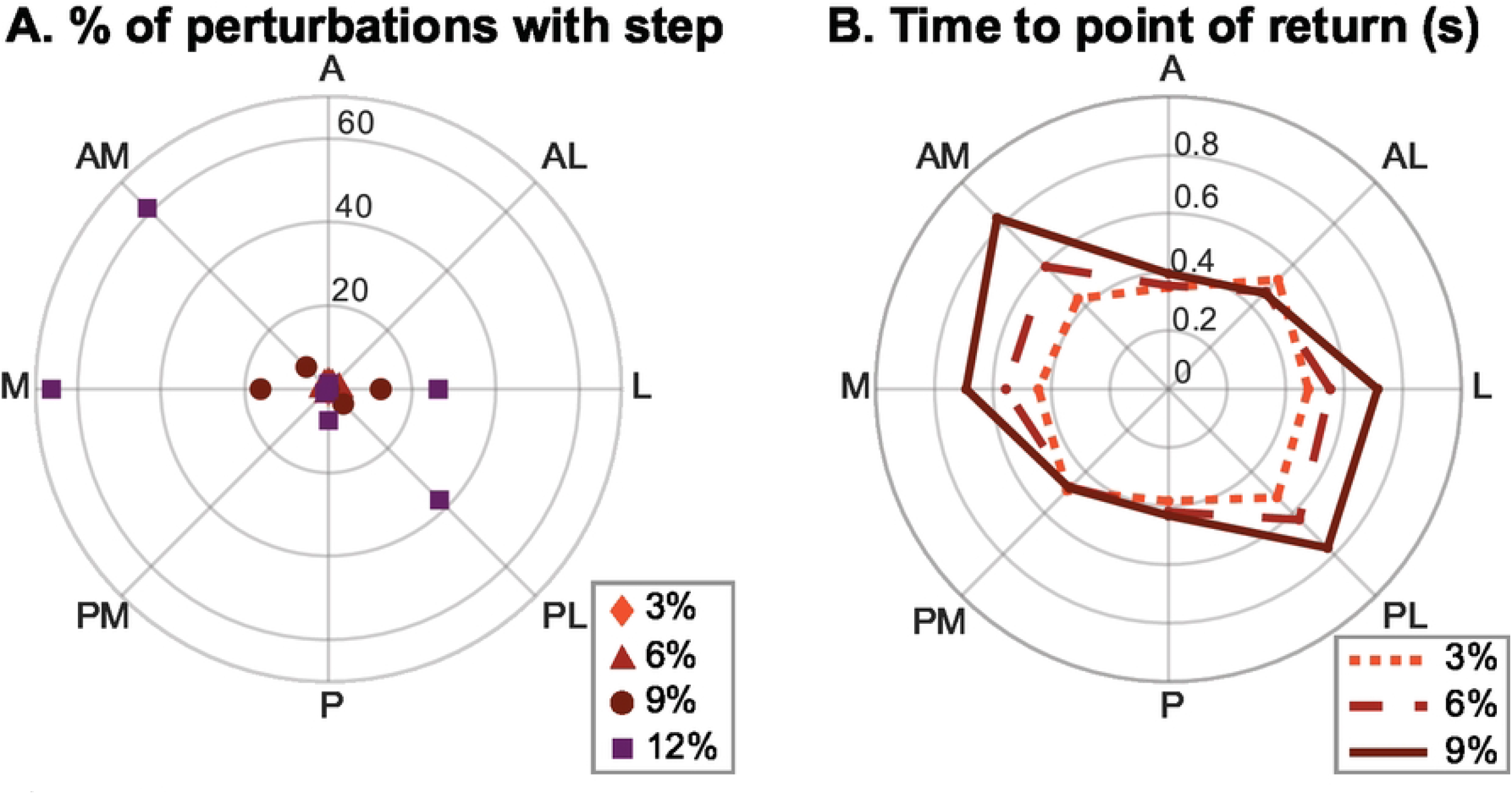
Effect perturbation on stepping and time to point of return. All based on group averages. A) The percentage of the perturbations in each direction and magnitude after which a step was required. B) Time needed to reach the point of return for each perturbation magnitude and direction.

The perturbations induced a motion of the CoM in the direction of the perturbation, Fig 3. If the perturbations were transverse to the line between both feet the total CoM deviations were larger compared to those after perturbations collinear with the line between both feet. For almost all perturbation directions, increasing the perturbation magnitude significantly effected the CoM deviation in both the ML and AP direction (detailed outcomes in the form of time series and complete results of the statistical tests can be found in the supplementary material, S1 Fig and S1 Table). This means that the perturbation also affects the plane perpendicular to the perturbation plane. This was the case except for A perturbations, after which the CoM deviation did not significantly change in ML direction. Especially for perturbations in ML direction, a clear response was present in the sagittal plane in terms of CoM and CoP positioning. After M perturbations the CoM was brought more forward, while after L perturbations the CoM was brought backward. Generally the CoP followed a trajectory surrounding the CoM, allowing to steer the CoM back to the starting position. After the perturbations collinear with the line between both feet the CoP trajectory made a large deviation, making use of the size of the BoS and enabling the quick return towards the initial condition as shown in Fig 2. For the perturbations transverse to the line between both feet the CoP trajectory was limited by the BoS boundaries.

**Fig 3.**
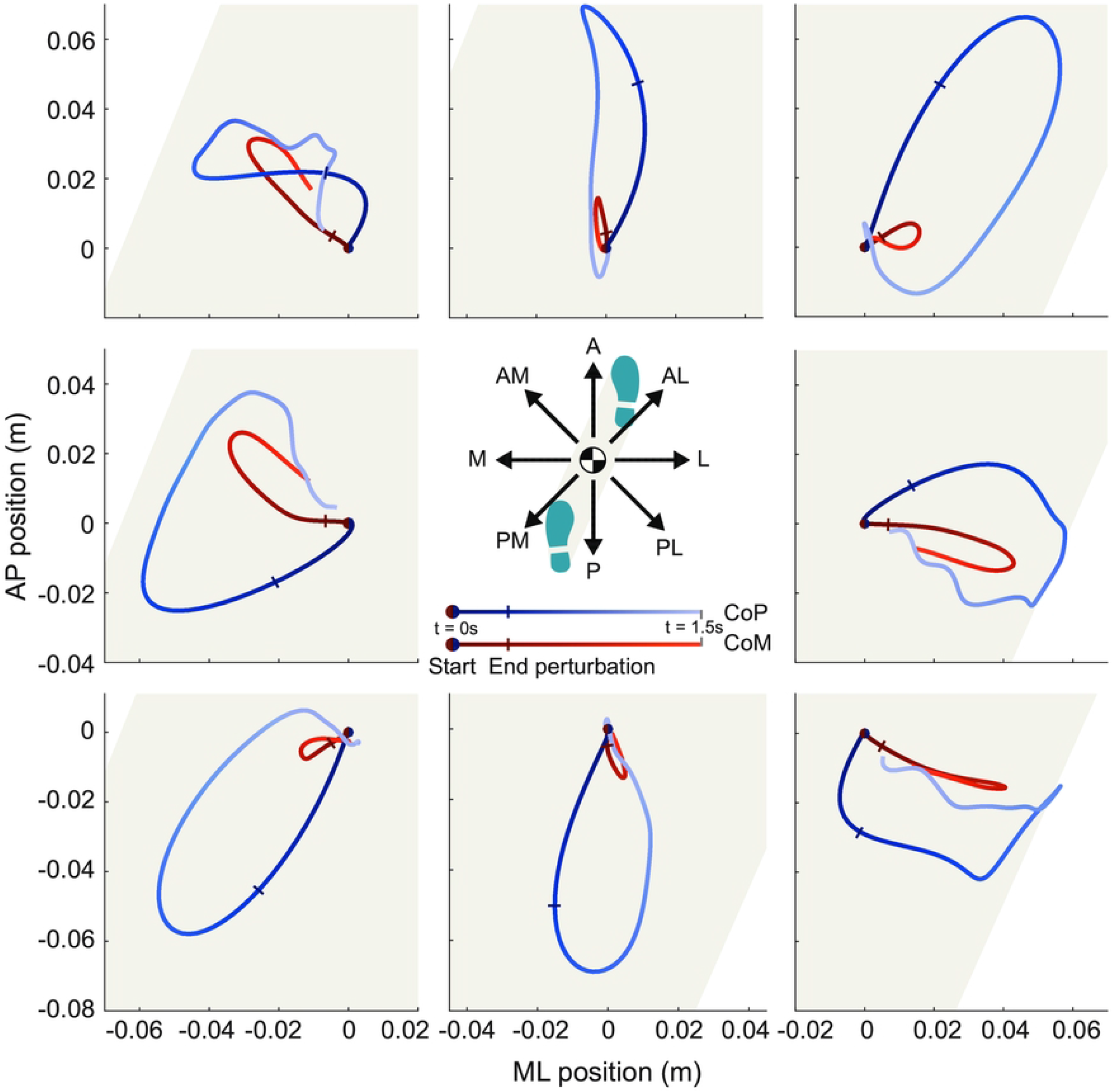
Top-down view CoM and CoP. Top-down view of the centre of mass (CoM in red) and centre of pressure (CoP in blue) trajectories after a 9% magnitude perturbation given in the direction indicated in the middle of the figure. A trajectory of 1.5 s is presented, with the start of the perturbation (t = 0 s) indicated with a dot and the end of the perturbation with a perpendicular line. The shaded area indicates the base of support. The presented results are the averages across all participants.

### Muscle level response

Due to the asymmetrical position of the feet in staggered stance, we observed different muscle responses in the trailing and leading leg to the same perturbations, Fig 4. The muscles acting around the ankle, the mSOL, mTA and mPL, exhibited a stronger response in the more loaded trailing leg compared to the leading leg. Conversely, the muscles around the hip, the mGMa and mGMe of the leading leg showed more prominent activations compared to those of the trailing leg. Overall, the upper and lower leg muscles of both legs showed a minimal response to perturbations in the A and P direction, compared to other directions. The perturbations applied in the directions transverse to the line between both feet elicited the largest reactions in all measured muscles.

**Fig 4.**
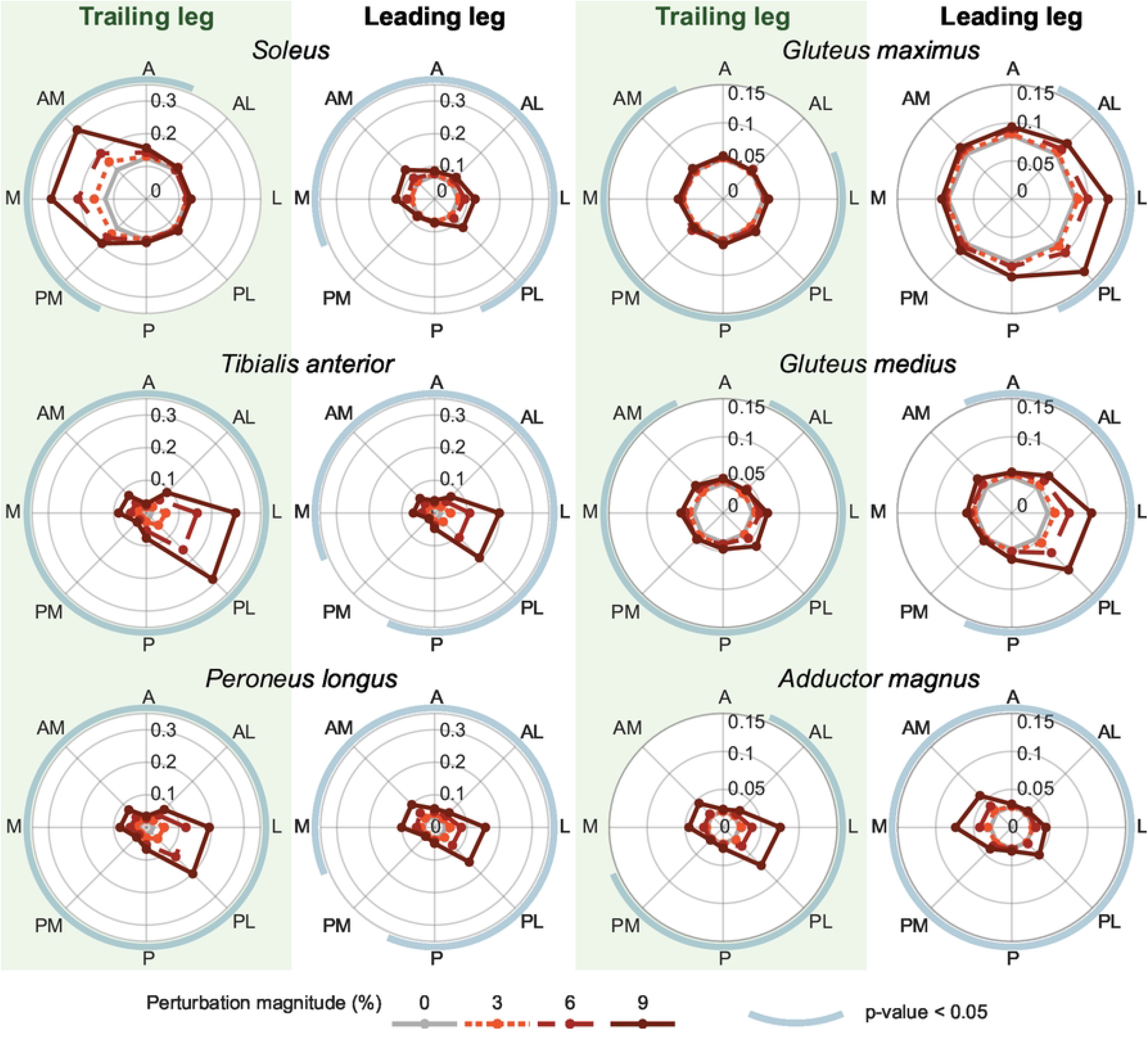
Normalised muscle activity. Muscle activations over the first 1.5 s after the perturbations given in the different directions. The results are presented for the trailing (= left) leg and leading (= right) leg. The line colour and style indicate the perturbation magnitudes. The blue circular arcs around the polar plots indicate whether there is a significant effect of the perturbation on the outcome measure with p *<* 0.05. All results are based on the averages across all participants.

For both legs the mPL, inducing plantarflexion and eversion and the mTA inducing dorsiflexion of the ankles, showed significant activations after perturbations in almost all directions, with the most prominent response seen after L and PL perturbations. The mSOL, inducing plantarflexion, exhibited a larger difference between the trailing and leading leg. The trailing leg mSOL had the largest response to predominantly M and AM perturbations. While the mSOL of the leading leg had a much smaller response but significant for a larger range of perturbation directions.

Within the same leg, the mGMe, a hip abductor, and mGMa, a hip extensor, showed similar reactions for the same perturbation directions. The gluteus muscles of the leading leg showed large significant activation patterns after perturbations in the L and PL directions. In comparison, the gluteus muscles of the trailing leg showed smaller responses overall, but they were significant for almost all directions except the A direction. Hip adductor mAM of both legs showed an opposite reaction to the same perturbations, with the strongest activations after perturbations transverse to the line between both feet.

### Joint moment response

The strongest and most significant joint contributions were observed after perturbations transverse to line between both feet, Fig 5. The lumbar joint mainly contributed in the frontal plane by creating a bending moment, while hardly any significant contributions were shown in the sagittal plane by lumbar extension. In the frontal plane the strongest joint contributions came from the loaded trailing leg. The hip ab- and adduction moments showed opposite responses in the in- and decrease of the abduction moment for the trailing and leading leg, contributing to bringing the upper body back to the starting position in ML direction. This was done together with an in- or decrease of the ankle inversion of the trailing foot after M and AM or L and PL perturbations respectively. Meanwhile there was no contribution of the ankle in- and eversion of the leading leg.

**Fig 5.**
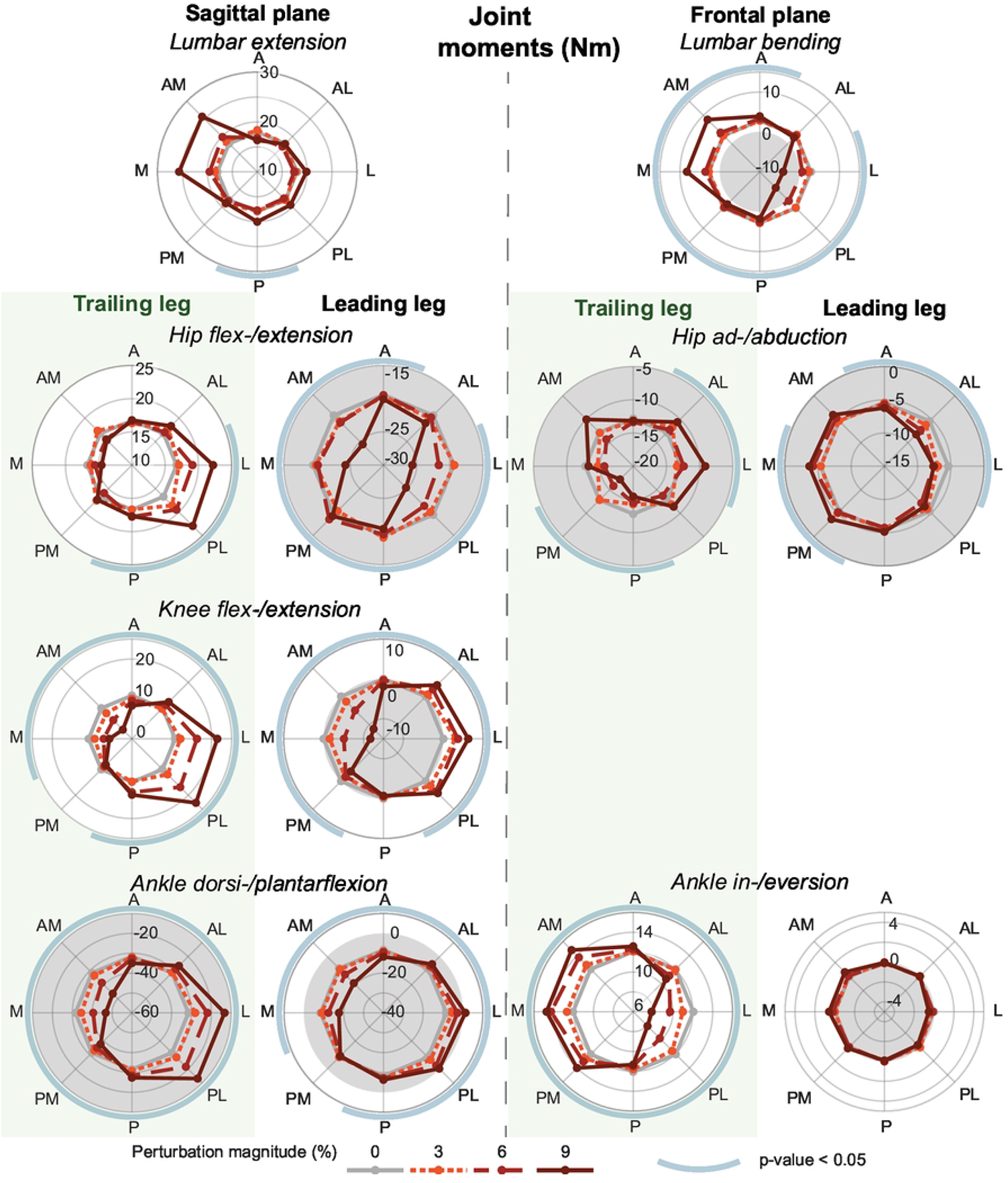
Joint moments. Joint moments of the lumbar, hip, knee and ankle joint in the frontal and sagittal plane in Nm. The mean value over the first 1.5 s, is shown for each perturbation magnitude and direction and the baseline during standing in the staggered stance posture. The line colour and style indicate the perturbation magnitudes. The blue circular arcs around the polar plots indicate whether there is a significant effect of the perturbation on the outcome measure with p *<* 0.05. The gray background indicates a negative value. All results are based on the averages across all participants.

In the sagittal plane the ankle, knee and hip joints of both the trailing and leading leg significantly contributed to the recovery after various perturbation directions. Perturbations transverse to the line between both feet strongly increased the hip extension moment of the leading leg. For the trailing leg a strong increase of the hip flexion moment was only seen after perturbations in L and PL directions. Symmetric responses in the change of the flexion/extension and plantar/dorsiflexion moments were seen in the knees and ankles respectively of both the trailing and leading leg. Especially after perturbations transverse to the line between both feet a strong increase (after M and AM perturbations) and reduction (after L and PL perturbations) of the ankle plantar flexion moment were presented.

## Discussion

This study aimed to establish how activations of the hip, knee and ankle joints in multiple directions contribute and co-operate in balance recovery after pelvis perturbations in various directions while standing in a staggered stance. As reported before by others, the effect of the perturbations on the maintenance of balance was clearly influenced by the dimensions of the BoS [4, 6, 11]. While standing in a staggered stance this made the individuals more sensitive to perturbations given in the ML and diagonal directions, transverse to the line between both feet. This higher sensitivity was reflected in a larger number of steps that needed to be taken, a longer time to the point of return, larger deviations of the CoM and stronger responses on muscle and joint level, compared to perturbations in line with the BoS.

After pure M or L perturbations, the CoM did not only deviate in ML direction, but also a significant AP motion was present. Remarkably, instead of moving the CoM away from the edges of the BoS, the CoM was brought more forward after the M perturbations and backward after the L perturbations, bringing the CoM closer to the BoS boundaries. Besides the fact that this might feel counter-intuitive, it was also contrary to what initially could be expected based on the presented ankle plantar-/dorsiflexion moments and contributing muscle activity. However, an explanation for this resulting CoM motion could be that it does assist in bringing the weight distribution back to the original situation [26]. After the M perturbations the ankle response resembled the ankle push-off during gait reported by Kim et al. [27]. Based on simulations they showed how the ankle push-off can contribute to ML balance during walking [27]. The combination of hip and knee flexion and extension moments contributed to achieving the presented CoM motion, which seems opposite to the findings of Winter et al. [4], during unperturbed staggered stance. They reported a counteraction between the ankle and hip as well, however the other way around, such that the ankle had to compensate for an inappropriate hip contribution.

Another remarkable finding after perturbations in L, PL and AM direction are the oscillations in the CoP trajectory. While checking the individual repetitions, oscillations were present as well, together with a larger variety of shapes of the CoP trajectories. The reason these perturbations resulted in these oscillations and larger variety, could be because the total CoP position is largely dependent on the loading and unloading of both legs [28]. If a perturbation brings the CoM close to the edge of the BoS, the participant might be close to the point that a step is needed for balance recovery. Preparations for making a step involve a weight shift towards the future stance leg. However if this is not needed in the end, it could result in a fast alternating weight shift between both feet.

In general, perturbations collinear with the line between both feet provoked smaller responses on joint level, compared to those transverse to the line between both feet. At the same time the excursion of the CoP was large, keeping the CoM deviations small. Especially responses in the frontal plane were small and not always significant after the perturbations collinear with the line between both feet, suggesting a weaker coupling between the sagittal and frontal plane compared to the perturbations transverse to the line between both feet. O’Connor et al. [11] extensively reported sensitivities after visual ML and AP perturbations during walking, normal stance and tandem stance. Our findings, while standing in a posture in between normal and tandem stance, revealed sensitivities similar to those reported by O’Connor et al. during walking and tandem stance [11]. This is probably because the shape and direction of the BoS during these postures correspond most with those during staggered stance.

Our outcomes might have been affected by the fact that standing in a staggered stance reflecting double support is not a posture people would often take naturally. Therefore firstly the opposed posture could have felt uncomfortable for the participants and might have affected the observed responses. Secondly, all perturbations were only applied with the right foot leading. Recommendations for future research would be to investigate the balance response in a staggered stance with the left leg leading as well, since the balance response of an individual could be influenced by the position of their dominant foot [5, 28]. Lastly, the participants were asked to cross their arms over their abdomen to prevent them from using arm swing as balance strategy. Swinging the arms and grabbing the adjacent rails as a reflex could assist in a natural balance response. This may have led to a psychological influence on the necessity of stepping. Besides, with the used setup it was also not possible to leave the arms along the body because of the rod connection with the motor on the side.

The obtained results give insights in balance recovery during staggered stance, a posture which becomes important while walking very slowly. Even-though the staggered stance posture might not be the most natural position, there are scenarios in which it becomes important to maintain balance in this posture. For example when walking very slowly or while wearing a lower limb exoskeleton due to a movement disorder. Therefore the results of this study provide fundamental insights into balance properties and abilities in this posture. This could facilitate improvement of future designs of exoskeleton controllers that assist paraplegics during walking.

## Conclusion

While standing in staggered stance, pelvis perturbations transverse to the line between both feet required strong joint responses in order to maintain balance. Reactions in both the frontal and sagittal planes contributed to the recovery of these perturbations. In contrast, perturbations collinear with the line between both feet revealed smaller responses and less coupling between responses in the sagittal and frontal plane.

## Data Availability

The data used for this study is published on 4TU.ResearchData and can be found via the following DOI: http://dx.doi.org/10.4121/20318382

## Supporting information

**S1 Fig. Time series results**. File containing the time series over the first 1.5 s after the perturbations for the various outcome measures after the different perturbation directions and magnitudes.

**S1 Table. Statistical results**. File containing tables with the results of all statistical tests.

## Acknowledgments

This work is part of the research program Wearable Robotics with project number P16-05, which is (partly) funded by the Dutch Research Council (NWO).

## References

1. Gorgey AS, Sumrell R, Goetz LL. Exoskeletal Assisted Rehabilitation After Spinal Cord Injury. Fifth edit ed. Elsevier Inc.; 2019.

2. Ren Z, Deng C, Zhao K, Li Z. The development of a high-speed lower-limb robotic exoskeleton. Science China Information Sciences. 2018;62(62):1–3. doi:10.1007/s11432-018-9717-2.

3. Bruijn SM, van Dieën JH, Meijer OG, Beek PJ. Is slow walking more stable? Journal of Biomechanics. 2009;42(42):1506–1512. doi:10.1016/j.jbiomech.2009.03.047.

4. Winter DA, Prince F, Frank JS, Powell C, Zabjek KF. Unified theory regarding A/P and M/L balance in quiet stance. Journal of Neurophysiology. 1996;75(75):2334–2343. doi:10.1152/jn.1996.75.6.2334.

5. Wang Z, Jordan K, Newell KM. Coordination Patterns of Foot Dynamics in the Control of Upright Standing. Motor Control. 2012;16:425–443.

6. Hof AL. The equations of motion for a standing human reveal three mechanisms for balance. Journal of Biomechanics. 2007;40(40):451–457. doi:10.1016/j.jbiomech.2005.12.016.

7. van Mierlo M, Vlutters M, van Asseldonk EHF, van der Kooij H. Centre of pressure modulations in double support effectively couneract anteroposterior perturbations during gait. Journal of Biomechanics. 2021;126. doi:10.1016/j.jbiomech.2021.110637

8. Horak FB, Nashner LM. Central programming of postural movements: adaptation to altered support-surface configurations. Journal of Neurophysiology. 1986;55(55):1369–1381. doi:10.1152/jn.1986.55.6.1369.

9. van Mierlo M, Ambrosius JI, Vlutters M, van Asseldonk EHF, van der Kooij H. Recovery from sagittal-plane whole body angular momentum perturbations during walking. Journal of Biomechanics. 2022;141. doi:10.1016/j.jbiomech.2022.111169

10. Wang Z, Newell KM. Asymmetry of foot position and weight distribution channels the inter-leg coordination dynamics of standing. Experimental Brain Research. 2012;222(222):333–344.

11. O’Connor SM, Kuo AD. Direction-dependent control of balance during walking and standing. Journal of Neurophysiology. 2009;102(102):1411–1419. doi:10.1152/jn.00131.2009.

12. Rietdyk S, Patla AE, Winter DA, Ishac MG, Little CE. Balance recovery from medio-lateral perturbations of the upper body during standing. Journal of Biomechanics. 1999;32:1149–1158.

13. Henry SM, Fung J, Horak FB. EMG responses to maintain stance during multidirectional surface translations. Journal of Neurophysiology. 1998;80(80):1939–1950. doi:10.1152/jn.1998.80.4.1939.

14. Nashner LM, Woollacott M, Tuma G. Organization of rapid responses to postural and locomotor-like perturbations of standing man. Experimental Brain Research. 1979;36(36):463–476. doi:10.1007/BF00238516.

15. Runge CF, Shupert CL, Horak FB, Zajac FE. Ankle and hip postural strategies defined by joint torques. Gait and posture. 1999;10:161–170.

16. Matjačić Z, Voigt M, Popović D, Sinkjær T. Functional postural responses after perturbations in multiple directions in a standing man: a principle of decoupled control. Journal of Biomechanics. 2001;34(34):187–196. doi:10.1016/S0021-9290(00)00182-2.

17. Mihelj M, Matjačić Z, Bajd T. Postural activity of constrained subject in response to disturbance in sagittal plane. Gait and Posture. 2000;12(12):94–104. doi:10.1016/S0966-6362(00)00065-5.

18. Mille M, Rogers MW, Martinez K, Hedman LD, Johnson ME, Lord SR. Thresholds for Inducing Protective Stepping Responses to External Perturbations of Human Standing. Journal of Neurophysiology. 2003;90:666–674.

19. Rietdyk S. Postural responses to unexpected multi directional upper body perturbations; Thesis, 1999.

20. Yoshida Y, Takeuchi K, Miyamoto Y, Sato D, Nenchev D. Postural balance strategies in response to disturbances in the frontal plane and their implementation with a humanoid robot. IEEE Transactions on Systems, Man, and Cybernetics: Systems. 2014;44(44):692–704. doi:10.1109/TSMC.2013.2272612.

21. Lee YJ, Aruin AS. Three components of postural control associated with pushing in symmetrical and asymmetrical stance. Experimental Brain Research. 2013;228(228):341–351. doi:10.1007/s00221-013-3567-4.

22. van der Kooij H, Fricke SS, van ‘t Veld RC, Prieto AV, Keemink AQL, Schouten AC, et al. Identification of hip and knee joint impedance during the swing phase of walking. IEEE transactions on neural systems and rehabilitation engineering. 2022;30.

23. Aggarwal A. SENIAM: Surface ElectroMyoGraphy for the Non-Invasive Assessment of Muscles; 2013. http://www.seniam.org/.

24. Wu AR, Simpson CS, van Asseldonk EHF, van der Kooij H, Ijspeert AJ. Mechanics of very slow human walking. Scientific Reports. 2019;9(9):1–10. doi:10.1038/s41598-019-54271-2.

25. Delp SL, Anderson FC, Arnold AS, Loan P, Habib A, John CT, et al. OpenSim: Open-source software to create and analyze dynamic simulations of movement. IEEE Transactions on Biomedical Engineering. 2007;54(54):1940–1950. doi:10.1109/TBME.2007.901024.

26. Reiman H, Fettrow T, Jeka J. Strategies for control of balance during locomotion. Human Kinetics 2018;7(7):18-25. doi.org/10.1123/kr.2017-0053

27. Kim M, Collins, SH. Once-per-step control of ankle push-off work improves balance in a three-dimensional simulation of bipedal walking. IEEE transactions on robotics. 2017;33(33):406–418. doi.org/10.1109/TRO.2016.2636297

28. Winter DA. Human balance and posture control during standing and walking. Gait posture. 1995;3(3):193–214.

